# Characterising SARS-CoV-2 viral clearance kinetics to improve the design of antiviral pharmacometric studies

**DOI:** 10.1101/2021.01.06.21249368

**Authors:** James A Watson, Stephen Kissler, Nicholas PJ Day, Yonatan Grad, Nicholas J White

## Abstract

A consensus methodology for pharmacometric assessment of candidate SARS-CoV-2 antiviral drugs would be useful for comparing trial results and improving trial design. The time to viral clearance, assessed by serial qPCR of nasopharyngeal swab samples, has been the most widely reported measure of virological response in clinical trials, but it has not been compared formally with other metrics, notably model-based estimates of the rate of viral clearance. We analysed prospectively gathered viral clearance profiles from 280 infection episodes in vaccinated and unvaccinated individuals. We fitted different phenomenological pharmacodynamic models (single exponential decay, bi-exponential, penalised splines) and found that the clearance rate, estimated from a mixed effects single exponential decay model, is a robust pharmacodynamic summary of viral clearance. The rate of viral clearance, estimated from viral densities during the first week following peak viral load, provides increased statistical power (reduced type 2 error) compared with time to clearance. We recommend that pharmacometric antiviral assessments should be conducted in early illness with serial qPCR samples taken over one week.

## Introduction

Acute SARS-CoV-2 infection can be characterised approximately as two overlapping clinical stages. The first pre-symptomatic stage comprises uncontrolled viral replication. Peak viral loads in the nasopharynx or oropharynx of individuals with symptomatic COVID-19 illness occur around the time of symptom onset [1]. The second stage of infection comprises a first order decrease in the viral load resulting from activation of host-defence mechanisms. Viral multiplication is attenuated, and clearance is augmented by effective host defences [2]. During this second stage a small subset of infected individuals (*<*5%) progress to severe pneumonia and some will die. The risk is strongly age dependent [3] and it is reduced substantially by vaccination [4]. Infections with the now-prevalent Omicron variant are associated with a lower risk of hospitalisation.

Effective antiviral drugs or biologics attenuate viral multiplication [5, 6]. Antiviral interventions are most effective early in the course of disease, while immunomodulators are life-saving in hospitalised patients as immunopathology dominates after approximately one week of illness [7]. Effective antiviral medicines administered early during SARS-CoV-2 infections should reduce the overall viral load, accelerate virus clearance, and reduce the probability of progression to severe COVID-19 illness. This has been shown most convincingly for the monoclonal antibody mixture of casirivimab and imdevimab (REGN-CoV-2) [5], and recently for the ribonucleoside analogue molnupiravir [6, 8].

Primary end-points in hospital based randomized controlled trials (RCTs) have usually been either the need for respiratory support or death whereas, in outpatient studies, measures such as changes in symptom severity or evidence of clinical progression (e.g. SpO_2_ *<*92%) are used generally [9, 10, 11]. Large sample sizes are required to detect meaningful effects in outpatient studies, even if high risk subgroups only are enrolled, as only a minority of symptomatic individuals develop hypoxaemia and progress to require hospitalisation [9]. In vaccinated populations progression to severe disease is now unusual. How then can the long list of repurposed candidate antivirals be prioritised for phase 3 evaluation, and how can the expanding number of candidate new antivirals be compared? Accurate measurement of viral clearance is potentially the most efficient method of ‘phase 2’ candidate drug screening and dose-finding, but there is currently no consensus on the pharmacometric methodology.

In the first week after becoming symptomatic with COVID-19, viral concentrations in nasopharyngeal or oropharyngeal swab samples decline exponentially [12, 13]. It is increasingly evident that the pattern of viral elimination is often bi-exponential, although the second phase, which is close to the limit of accurate viral quantitation, may be unobserved if follow-up is not extended. In the context of a clinical trial viral clearance can be summarised in many different ways. The most commonly employed is time-to-clearance (the time to reach the lower limit of detection for a given assay) [11] but this is dependent on both the assay used and the baseline viral load (i.e. the viral load at enrolment in the study, which will depend on how cases are ascertained and the individual peak viral loads which vary substantially across individuals). We used prospectively gathered data from 280 infection episodes from individuals who had frequent qPCR sampling before and after peak viral load to characterise the kinetics of viral clearance and to propose robust summary statistics as pharmacodynamic endpoints for pharmacometric evaluations and phase 2 type clinical trials.

## Materials and Methods

### Data

We obtained prospective longitudinal SARS-CoV-2 RT-qPCR testing data on 474 infection episodes from individuals in the US National Basketball Association’s (NBA) occupational health program, observed between November 28 2020 and January 10, 2022. These infection data have been described previously in detail by [2, 12, 14]. The data are openly accessible on github at https://github.com/gradlab/CtTrajectories_AllVariants and https://github.com/gradlab/CtTrajectories_Omicron. In brief, clinical samples were obtained by combined swabs of the anterior nares and oropharynx for each individual administered by a trained provider. Viral load was measured using the cycle threshold (CT) according to the Roche cobas target 1 assay. Vaccination information was reported and verified by NBA staff and the clinical operational team.

### Data pre-processing

For each infection episode, we initially defined day 0 as the day with the lowest observed CT value (i.e. highest nasopharyngeal or oropharyngeal viral loads). We selected all infection episodes where there were at least 5 samples with a CT value less than 40 and at least one sample with a CT value less than 30 in the time period -20 to +20 days for each individual (a total of 280 infections). To these infection data, we then fitted a simple Bayesian hierarchical ‘up-and-down’ phenomenological model to the ΔCT values (40-CT) [13]. The model was specified as:

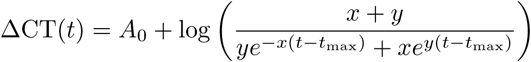

where *A*_0_ corresponds to the peak ΔCT occurring at time *t* = 0 (i.e. the intercept which is proportional to the peak log viral load); *x* is the growth rate in the initial viral proliferation stage; and *y* is the decay rate in the viral clearance stage. The main purpose of this model was to estimate the time of peak viral load (*t*_max_) for each individual. Individual random effect terms were specified for *A*_0_, *x, y* and *t*_max_ (all additive random effects). For each individual we then defined time zero as their mean estimated *t*_max_. We then selected all CT values taken during the interval [*t*_max_, *t*_max_ + 14]. This gave a total of 280 infection episodes with a median of 7 CT values post peak viral load measures per episode (range: 1 to 18).

### Model of viral clearance dynamics

To characterise the viral clearance stage in these 280 infection episodes, we fitted a series of phenomenological models. We chose not to fit mechanistic models (e.g. [15]) as the goal was not explain the biology underlying viral clearance but to provide a robust method from which to infer summary statistics for virological clinical trial endpoints.

#### Model 1: Exponential decay

The simplest model for viral clearance is log-linear decay specified as:

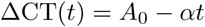

where *A*_0_ is proportional to the peak log viral load (intercept at *t* = 0) and *α* is the clearance rate.

#### Model 2: Bi-exponential decay

There is evidence of persistent viral shedding at low densities (CT values > 30) [12]. Bi-exponential decay (a “two-compartment” model) can fit this type of clearance profile, and is defined as:

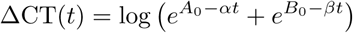

where *B*_0_ < *A*_0_ and *β* < *α*. Under this model, the intercept at time *t* = 0 is 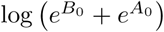 (which is approximately equal to *A*_0_ if *B*_0_ ≪ *A*_0_); *α* is the rate constant for the initial ‘fast’ decay; and *β* is the rate constant for the second stage slower decay.

#### Model 3: Regularised splines

We used penalised linear models from the R package *mgcv* which implements penalized regression splines with automatic smoothness estimation [16]. We specified random effect terms for each infection episode.

#### Error model for models 1-2

It was been observed previously that there is considerable variation in viral load estimates from oropharyngeal and nasopharyngeal swabs [12, 13]. This can result from several factors including variations in local viral densities, in swab size and swabbing technique, number of cells, volume of eluate, and qPCR assay variability. To account for these different sources of variation, in models 1-2 we specified the model likelihood as a mixture of two distributions (as in [13]). The first component was a Gaussian distribution with mean value the predicted ΔCT under the model (with left censoring at ΔCT=0) and variance *σ*_CT_. The variance *σ*_CT_ can be interpreted as the measurement noise under ‘normal’ conditions (i.e. well performed swab). The second component was a Gaussian distribution with fixed mean *μ*_noise_ and variance *σ*_noise_ which can be interpreted as the measurement noise in the ‘non-normal’ conditions, for example when the swab is badly performed. The mixing parameter *λ* determines how often the noise component occurs.

#### Hierarchical model and prior distributions

Models 1 and 2 were specified as hierarchical Bayesian models with random effect terms for the slope coefficient(s) and the intercept(s). For infection *i* for model 1 this was parameterised as:

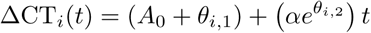

The first random effect *θ*_*i*,1_ represents an individual shift from the population intercept *A*_0_ (giving the individual log peak viral load); the second random effect *θ*_*i*,2_ represents the log of the proportional increase or decrease in the population rate constant *β*.

For model 2 the random effects were parameterised as:

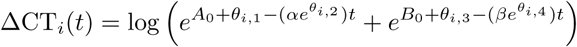

where *θ*_*i,k*_, *k* = 1..4 have analogous interpretations as in model 1.

We specified weakly informative priors for all parameters of the two Bayesian hierarchical models. The prior values were informed by previous analyses [12]:

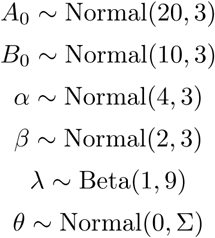

For *A*_0_ (the intercept on the ΔCT scale), the prior corresponds to a population mean nadir CT value between approximately 14 and 26. For *B*_0_ intercept this corresponds to the persistent viral shedding phase starting at around CT values of 30. The prior on *λ* indicates that roughly 1 in ten samples will have substantial error (i.e. be drawn from the error mixture component). The variance-covariance matrix Σ was parameterised as its Cholesky LKJ decomposition, where the L correlation matrix had a uniform prior (i.e. hyperparameter *ν*=1).

### Clinical trial simulation and power analysis

We used the posterior predictive distribution under model 2 (bi-exponential decay) to simulate clinical trial data under the assumption that patients were enrolled at exactly the time of their peak viral load. For each clinical trial simulation we drew a single sample from the posterior distribution of model 2, generated random effect terms for each individual in the trial, and then generated simulated CT data. We assumed that the clinical trial was comparing no intervention to an antiviral agent whose effect size *γ* was parameterised on the log scale as a constant scaling factor on the population mean rate coefficients *α* and *β*. We simulated trials that did twice daily swabs (performed independently) over either 7 or 5 days (a total of 16 or 12 viral load estimates). For each simulated dataset we then estimated the treatment effect by fitting the single exponential model (thus mis-specified by design). The treatment effect 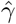 estimated under the log-linear model was also parameterised on the log scale representing the proportional change in the population clearance rate:

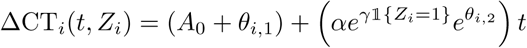

where *Z*_*i*_ is the randomised treatment allocation (1 is treated, 0 not treated) and 𝕝 is the indicator function taking value 1 if *Z*_*i*_ = 1 and 0 otherwise. We gave *γ* a Normal(0, 1) prior. If the 95% percentile interval for the posterior distribution over *γ* did not include 0, this was defined as rejecting the hypothesis that the intervention had no effect.

For comparison, we estimated Kaplan-Meier survival curves for the time to clearance in each treatment arm, whereby a clearance event was defined as both swabs on a given day being equal to 40 (ΔCT=0). We then tested for a difference between the survival curves using the log-rank test. Rejection of the null hypothesis was defined as a p-value <0.05. For both the rate of clearance and time to clearance simulations, the power was defined as the proportion of simulations which correctly rejected the null hypothesis of no treatment effect.

### Statistical analysis

All statistical analyses were performed in R version 4.0.2.

We defined the time to clearance as the time to the first recorded CT value equal to 40 with right censoring at the last recorded value if all values were less than 40. We compared the survival curves of the COVID-19 vaccinated individuals and the unvaccinated individuals using the log-rank test (as implemented in the R package *survival*).

For models 1-2 of viral clearance we considered CT values of 40 (ΔCT of 0) as left censored. The likelihood of the model parameters *θ* given the data is thus equal to 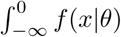, where *f*(·|θ) is the model likelihood.

Bayesian hierarchical models were coded in *stan* and fitted using Hamiltonian Monte Carlo with the *rstan* package [17]. For model 2, to avoid label switching across the parameters *A*_0_, *B*_0_ and *α, β*, we used the *ordered* vector type. Penalised additive linear models were fitted using the R package *mgcv* [16]. We checked convergence of chains by visual inspection of trace plots. All data and code are available at https://github.com/jwatowatson/SARS-CoV2-Viral-Clearance-Kinetics.

## Results

### Phenomenological models of viral clearance

After data pre-processing we had serial viral load data in 280 infection episodes after the inferred peak viral load with a median of 7 samples per infection (Figure 1). We fitted three phenomenological models to these post-peak clearance data: a single rate exponential decay Bayesian hierarchical model; a bi-exponential decay Bayesian hierarchical model; and a random effects additive linear model (penalised spline regression). All three models showed substantial bias in the distribution of the residuals as a function of the observed CT values, with the strongest bias appearing for the single exponential decay model. As the overall profile of viral clearance is clearly non-linear on the CT scale (as demonstrated by the daily median CT values shown by the pink triangles in Figure 1, note that the median is unbiased with left censoring), the linear model gives a compromise fit with a lower intercept and shallower slope. The bi-exponential decay model also had a biased residual distribution but the fit was substantially better for some infections which clearly demonstrated a second phase (see individual fits in supplementary Figures S1-S5). The penalised spline regression model did not give an improved fit compared to the bi-exponential model with respect to the residual distributions.

**Figure 1:**
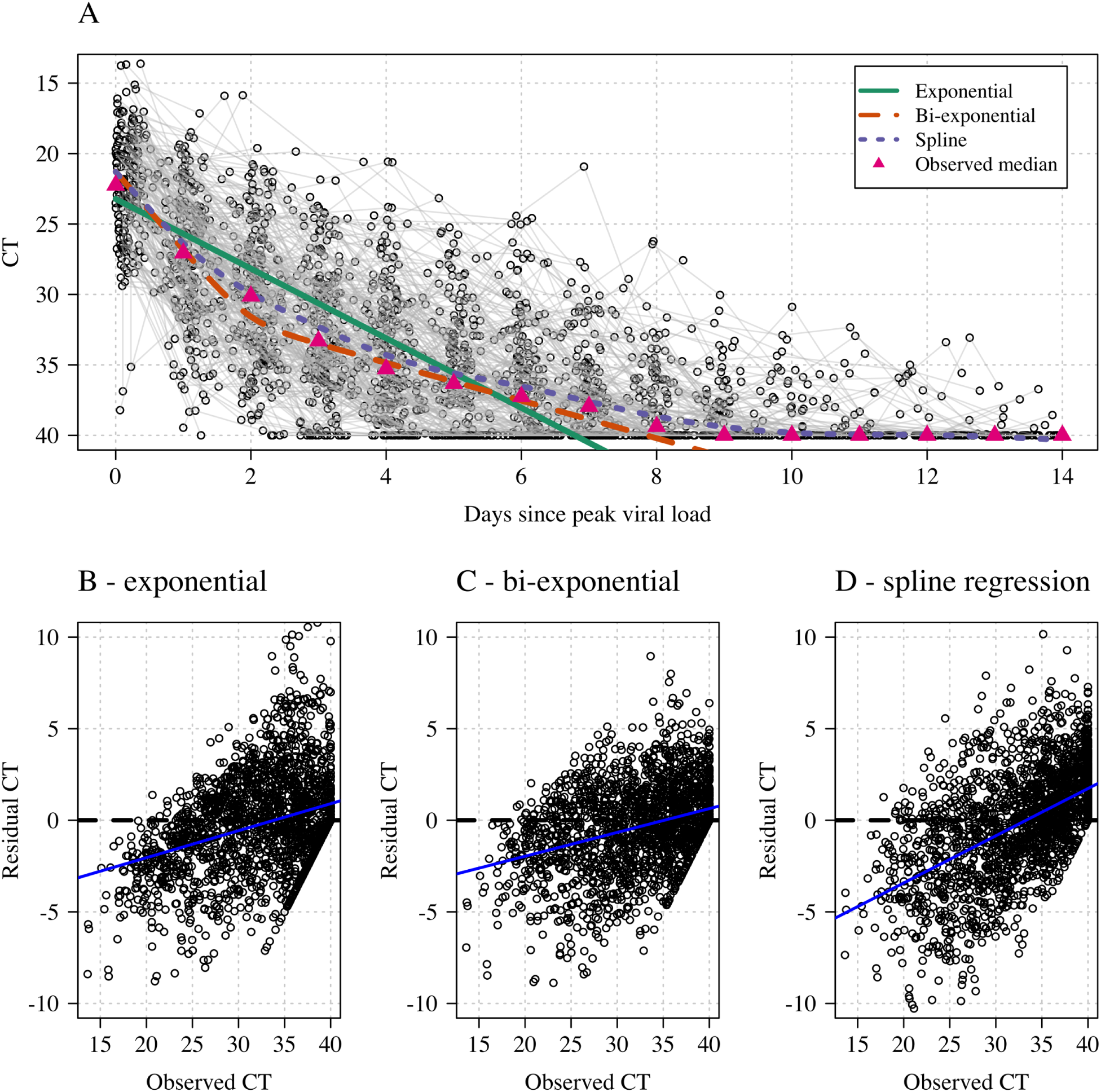
Comparing model fits for three phenomenological models of SARS-CoV-2 viral clearance. Panel A shows a spaghetti plot of the data with the mean predicted values from the three models (exponential: green; bi-exponential: orange; penalised splines: purple). The daily median CT values are shown by the pink triangles. Panels B-D show the distribution of the residuals as a function of the observed CT values (the blue lines show least-squares fits).

The bi-exponential model estimated a lower variance around the predicted mean value and a lower proportion of datapoints coming from the ‘noise’ component (supplementary Figure S6). Under the single exponential model approximately 8% of observations were fit to the noise component, whereas this proportion was only 2% for the bi-exponential decay model, indicating a much better fit overall.

### Pharmacodynamic summaries of viral clearance

For a single exponential decay model (model 1), viral clearance of the *i*^th^ infection can be summarised by the estimated individual clearance rate 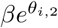. However, for a bi-exponential decay model (model 2), there are two individual clearance rates 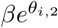 and 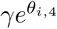 and an additional ‘elbow’ intercept *B*_0_ + *θ*_*i*,3_, thus giving three partial summaries of the viral clearance. These can be combined into a single metric by taking the area under the curve (AUC) of the estimated ΔCT_*i*_ with left censoring at 0. In order for the AUC to be independent of the baseline viral load, it is necessary to scale the AUC by the estimated intercept ΔCT_*i*_(*t* = 0). Note that for the single exponential model the AUC is directly proportional to the clearance rate.

Out of the 280 infection episodes, 77 had a confirmed vaccination status whereby 17 infections occurred in fully vaccinated individuals and 60 in non-vaccinated individuals. Consistent with the previously published analysis of the same dataset [2] and other separate data [13], the single exponential model suggested a faster clearance rate in the vaccinated compared with the unvaccinated subjects (p=0.05 comparing random effect terms for the slope coefficient *α*). As measured by the AUC estimated until day 5, day 7, or day 14, the log-linear model estimated larger reductions in AUC in vaccinated relative to unvaccinated individuals than the bi-exponential or the spline regression models (Figure 2).

**Figure 2:**
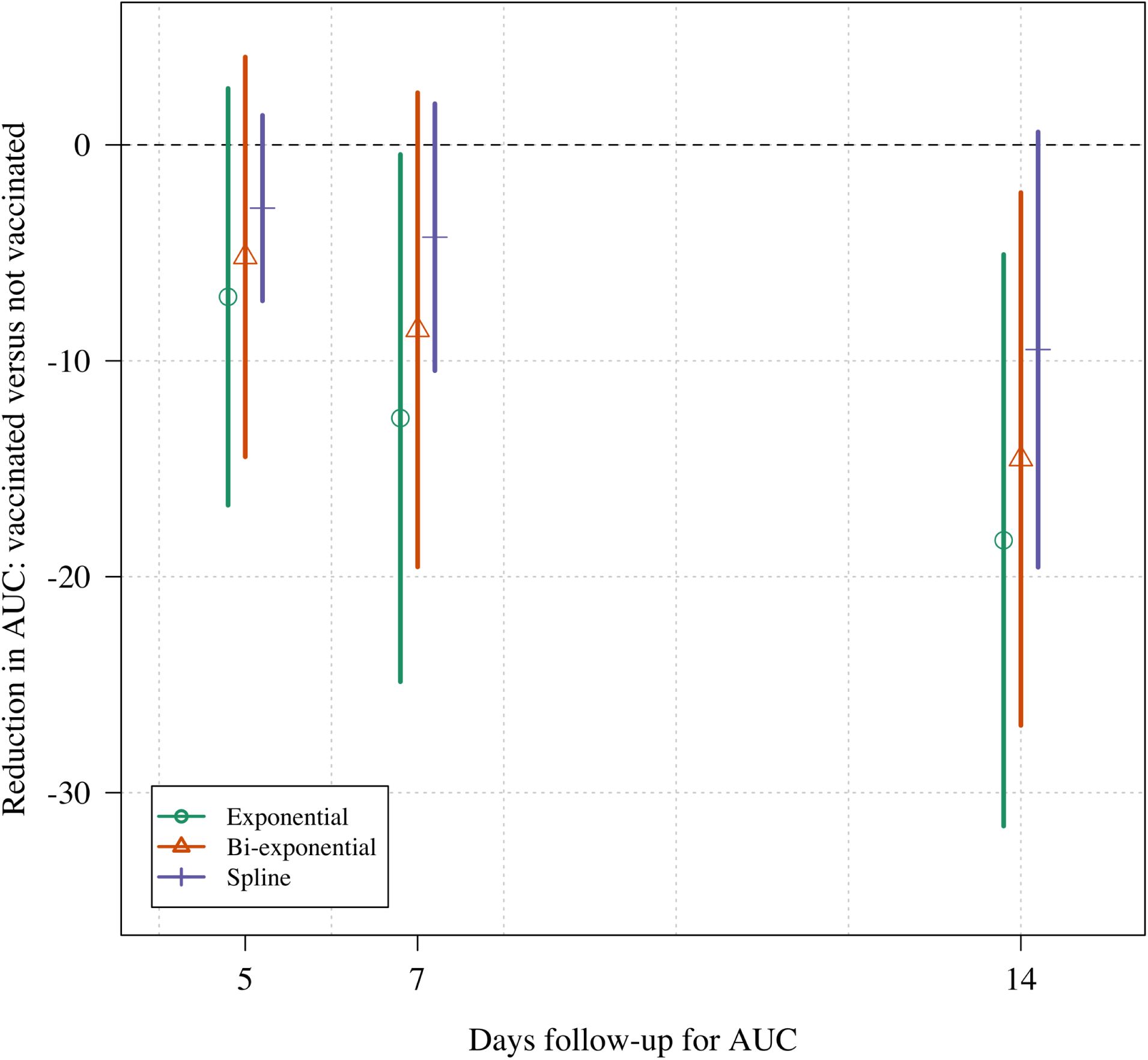
Difference in AUC for the estimated ΔCT in infection episodes in vaccinated (n=17) and unvaccinated (n=60) individuals. The estimated AUC is scaled by the intercept to remove the dependence on the baseline viral load. Approximate confidence intervals are calculated from a t-test.

The virological endpoint most commonly used in COVID-19 clinical trials is the time to viral clearance (a model-free measure, although the exact definition of this measure varies across trials). Time to clearance as a summary statistic is ‘inefficient’ because it is dependent on the initial viral load; it does not borrow information across time points; and it only uses the noisy estimate of when the viral load first reaches the lower limit of detection. It is also influenced by the variable presence of the second slower phase of viral clearance. There was no difference between time to clearance in the vaccinated versus unvaccinated individuals (Figure 3; p=0.2 for a log-rank test).

**Figure 3:**
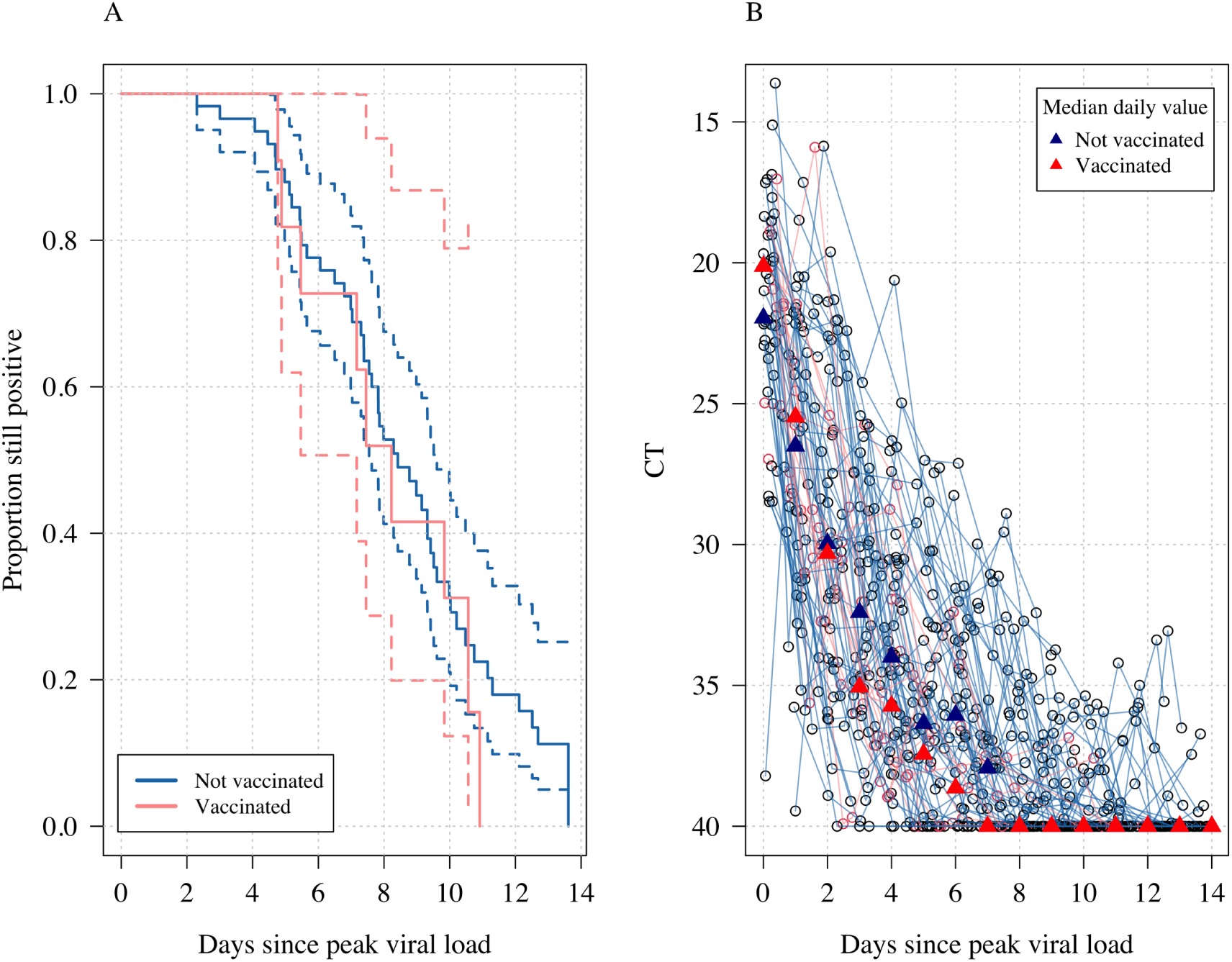
Time to viral clearance in vaccinated and unvaccinated individuals. Panel A shows the Kaplan-Meier survival curves (with 95% confidence intervals) of the proportion still testing positive over time. Panel B shows the individual viral load profiles with daily median values shown by the triangles. Pink: vaccinated; dark blue: unvaccinated.

### Sample size estimation

Previous work has estimated that sample sizes of approximately 500 patients per arm are required for clinical trials evaluating differences in time to viral clearance as primary end-points [11]. We postulated that by estimating the rate of clearance directly from daily serial viral load measurements, necessary sample sizes could be reduced substantially. By estimating the rate of clearance (even under a mis-specified model) information is borrowed across the serial viral load measurements, thereby resulting in a gain in efficiency relative to the standard approach of estimating the time to clearance. To explore this, we simulated data under the posterior predictive distribution of model 2 (bi-exponential decay) fitted to the NBA dataset. We assumed effect sizes equal to 30% and 50% increases in the population clearance rate parameters of the model. This results in approximately a median 3-4 cycle threshold difference by day 5. Figure 4A shows the median daily CT values over time for no treatment (black) and treatment with the two effect sizes tested.

**Figure 4:**
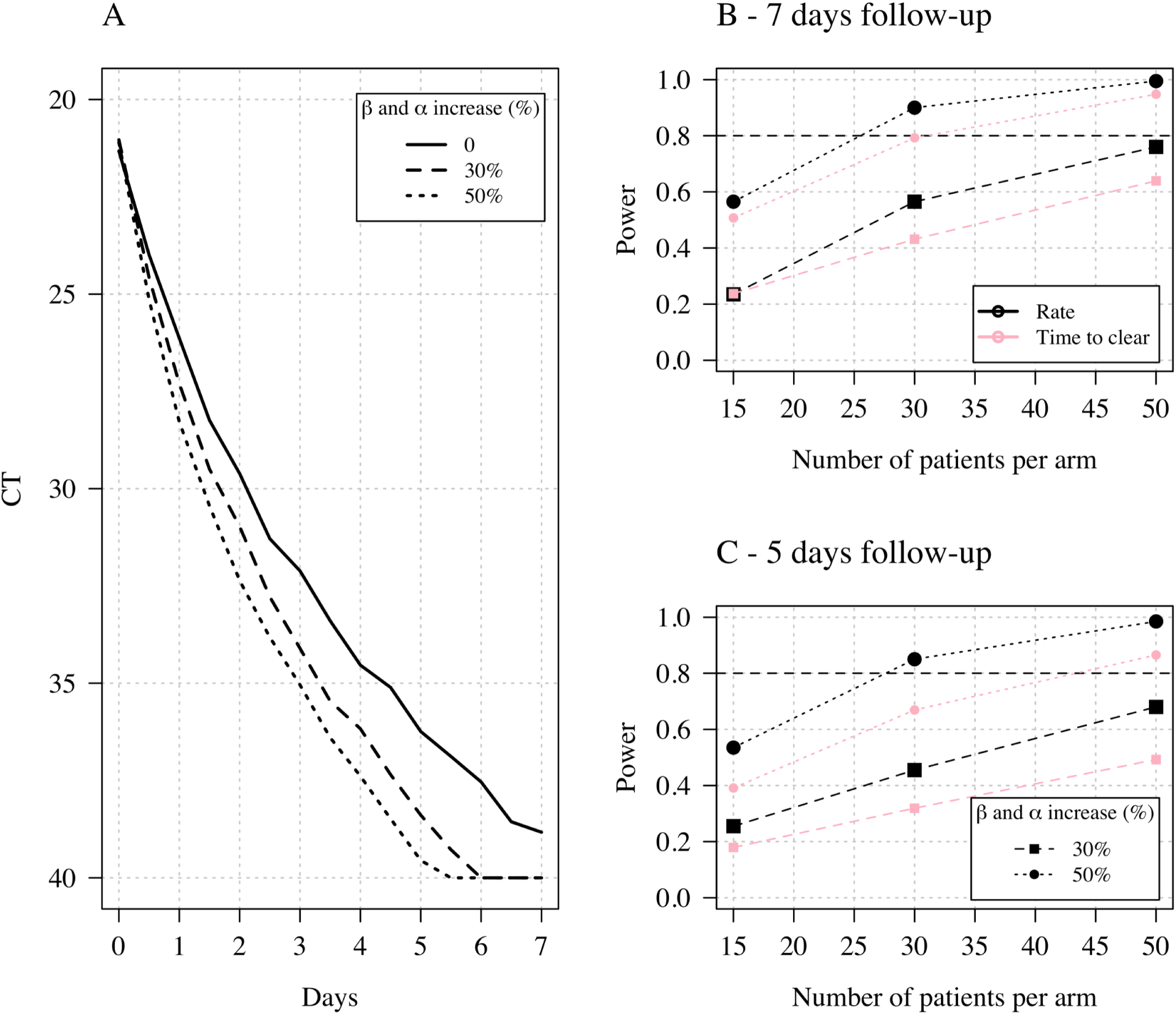
Power calculations based on data generated under a bi-exponential model for time to clearance or rate of clearance (single exponential model). Panel A shows the median CT values from the posterior predictive distribution under model 2 (bi-exponential decay) fit to the US NBA dataset. The thick line shows the median clearance profile under no intervention; the dashed and dotted lines show the median clearance profile for interventions with effect sizes of 30% and 50%, respectively (the effect size is defined as the proportional increase in rate coefficients *α, β* in the data generating model). Panels B and C show power estimations (1 - type 2 error) when comparing rates of clearance (black) under a single exponential model or time to clearance (pink).

We then estimated the power of a clinical trial with 1:1 randomisation between no treatment and an intervention with either of these treatment effects, whereby the null hypothesis of no treatment effect was either tested by modelling clearance rates directly (under a single exponential model, thus mis-specified by design) or by comparing survival curves for the time to clearance. Panels B & C of Figure 4 show the estimated power (1 - type 2 error) for sample sizes of 15, 30 or 50 patients per arm, with twice daily swabbing for either 7 or 5 days, respectively. In all simulations, greater power was obtained by modelling the rate of clearance directly even though this was done under a mis-specified model (time to clearance is a model free statistic). The average increase in power was approximately 10 percentage points.

## Discussion

An effective, well tolerated, safe, affordable and generally available treatment of COVID-19 that prevented progression to severe disease would be of enormous global health benefit. There have been many small clinical trials assessing the efficacy of repurposed candidate antivirals, justified usually by moderate inhibitory activity in virus cell cultures, but actionable evidence has come mainly from relatively few large randomised controlled trials in hospitalized patients. However, by the time patients require hospital care, viral loads have declined and immune pathology dominates the clinical picture. In these large RCTs immunomodulators (notably corticosteroids and IL-6 antagonists) have proved life-saving [7, 18]. These RCTs in hospitalised patients have not shown life-saving benefit from any of the antiviral repurposing candidates. The RNA-dependent RNA polymerase inhibitor remdesivir, developed originally to treat hepatitis C, was associated with a shorter duration of hospitalisation in a randomised controlled trial [19], and has received emergency use authorisations in many countries. But remdesivir did not reduce mortality in the large SOLIDARITY [18] and DisCoVeRy trials [20] and the WHO currently recommends against its use in COVID-19 [21].

Recently large RCTs in outpatients with uncomplicated COVID-19, mainly comprising “high risk” individuals (i.e older and with co-morbidities predisposing to more severe illness), have shown significant clinical benefits with both SARS-CoV-2 directed monoclonal antibodies [5] and small molecule antiviral drugs; notably the nucleoside analogue molnupiravir [6], and the ritonavir boosted viral protease inhibitor nirmatrelvir (as yet unpublished data). Early administration of remdesivir has also been shown to reduce hospitalisations [22]. Apart from these three, there are a large number of both repurposed and novel antiviral drugs either in development or under consideration for COVID-19 prophylaxis or treatment, but there is no agreed methodology for testing them in vivo or for determining the optimum dosage. It is simply not possible to conduct very large RCTs on each potential antiviral treatment, so a method of in-vivo pharmacometric assessment is needed in order to choose potential candidates for larger “phase 3” evaluations, to compare new antiviral therapies, and to optimise dosing. Although COVID-19 is a systemic infection, viral clearance from the upper respiratory tract is the only readily accessible measure of an ambulant patient’s virological response. Viral densities in nasopharyngeal or oropharyngeal swab samples peak around the time of clinical presentation and then exhibit a bi-exponential decline within the first week of illness. Previous vaccination or infection accelerates viral clearance [2, 13]. There is substantial variation between individuals, but mean profiles in untreated or placebo treated outpatients are very similar across large studies. With the notable exception of remdesivir [22], the antiviral medicines which have proved to be clinically effective in large RCTs do provide a significant acceleration of viral clearance.

Our analyses suggest that viral clearance is best assessed by measuring the approximate slope of the log-linear decline in qPCR densities (ΔCT values, accounting for left-censoring) over the first week after starting treatment. In many patients a bi-phasic elimination profile is evident, but the majority of viral load reduction is in the initial phase, and it is this phase which is accelerated by effective interventions (vaccination, monoclonal antibodies, small molecule antivirals). Although mis-specified, the log-linear model appears to capture in a robust way the overall trend of viral clearance. The widely reported time to viral clearance is a less precise measure, and it results in lower statistical power in pharmacometric studies. This is because the time to viral clearance depends on the frequency of testing, the sensitivity and reproducibility of the method and, critically, the initial virus density (and thus the stage of disease). It is also variably confounded by detection of the slower terminal virus elimination phase.

Viral clearance reflects both host-defence and any additional contribution from an antiviral therapeutic. Thus, the pattern of clearance will depend on the stage of disease and the host’s response. We propose using the slope of the initial log-linear decline in viral densities in nasopharyngeal or oropharyngeal secretions (i.e., the rate constant of the major decline) as the primary endpoint in phase 2 studies. The difference between the slopes with the putative antiviral and without represents the drug effect. Evaluations of therapeutic interventions which do not report these values and use time to viral clearance as the primary endpoint are difficult to interpret. Time-to-clearance results in a higher type 2 error (lower power) than rate-of-clearance, and therefore requires larger sample sizes, at least daily sampling, and longer duration of follow-up. It is therefore imprecise, inefficient, and consequently expensive. Importantly, any comparison or metaanalysis of studies which used different sampling techniques, or had different qPCR sensitivities, or different cut-offs, is confounded systematically if time-to-clearance is used as an endpoint, but not if rate-of-clearance is measured. It is important to note that comparisons between interventions should be contemporary as both immune status and viral variants are important contributors to viral clearance. Observational studies using historical controls would likely give misleading results.

There are several limitations to this study. The data were obtained from a monitoring scheme and not a prospective study of antiviral pharmacodynamics. The COVID-19 pandemic has evolved rapidly with an increasing proportion of the population being immunised and rapid changes in viral variants. More information on viral dynamics in infections with the Omicron variant in vaccinated individuals is needed. The rate constant of the decline in viral densities over one week is a hybrid parameter and more information is needed on virus clearance profiles in different settings to determine if this is the optimum pharmacodynamic measure in all circumstances. There is substantial variation in measured viral densities, even within an individual infection, and more information on the sources of variance is needed to refine these approaches.

In summary our model-based simulations suggest that pharmacometric assessment of candidate antivirals for COVID-19 should measure virus clearance rate, and not the much more widely used time to clearance, as their primary endpoint. If performed in early uncomplicated illness, reasonable precision can be obtained with twice daily qPCR samples taken over the course of one week after enrolment in each studied patient. Adaptive randomisation using the viral clearance rate can rapidly identify active intervention arms.

## Data Availability

All data and code are available on the github repositories linked to this manuscript.

https://github.com/gradlab/CtTrajectories

https://github.com/jwatowatson/phase2sims

## Acknowledgments

This research was funded, in whole or in part, by The Wellcome Trust. A CC BY or equivalent licence is applied to author accepted manuscript arising from this submission, in accordance with the grant’s open access conditions. NJW is a Principal Research Fellow funded by the Wellcome Trust (093956/Z/10/C). JAW is a Sir Henry Dale Fellow funded by the Wellcome Trust (223253/Z/21/Z).

## Supporting Information

**Figure S1:**
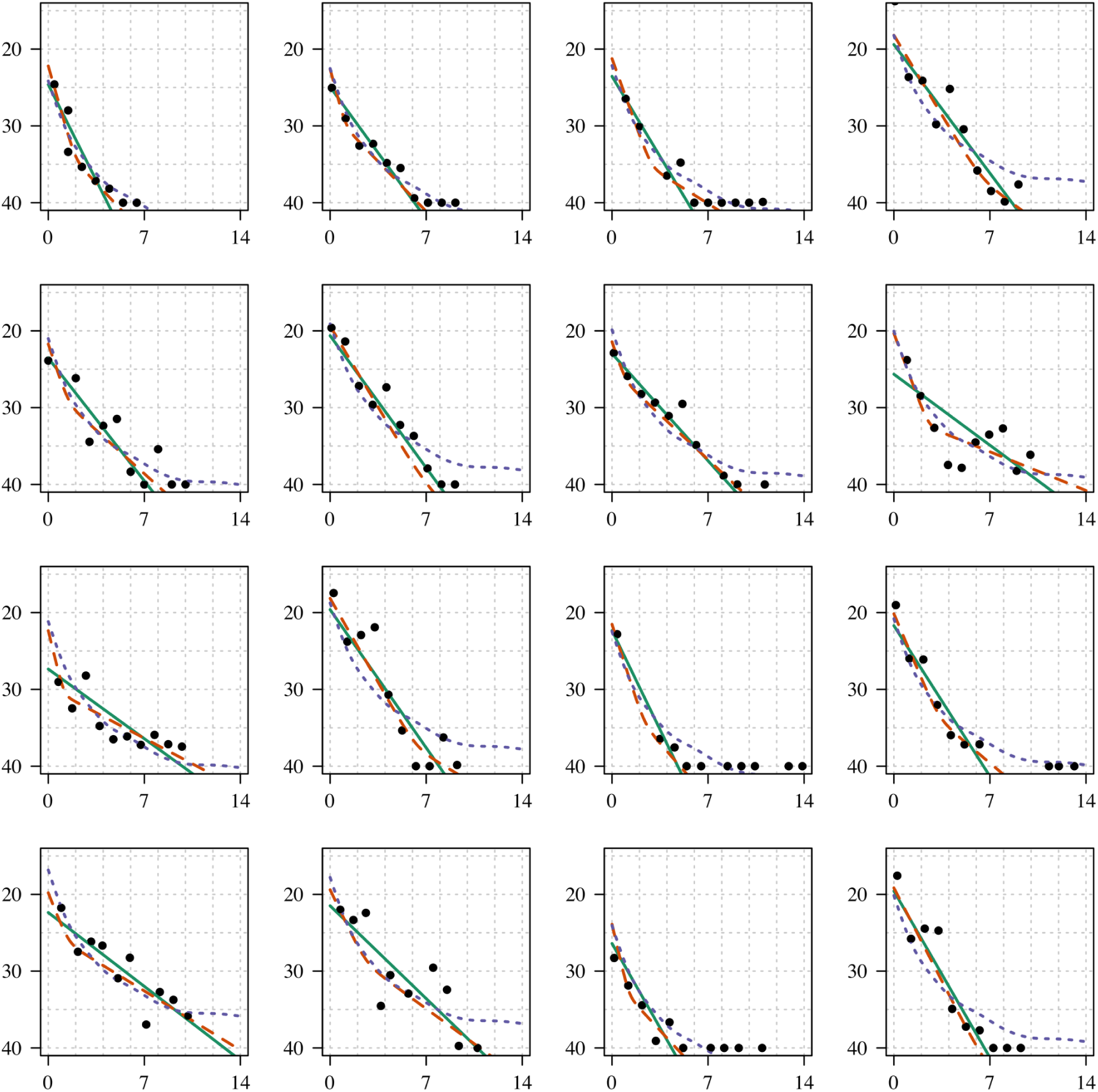
Individual model fits for infections with at least 10 CT values (panel 1). The x-axis is the time since peak viral load, the y-axis the CT value. The thick line shows the log-linear fit; the dashed line shows the bi-exponential fit.

**Figure S2:**
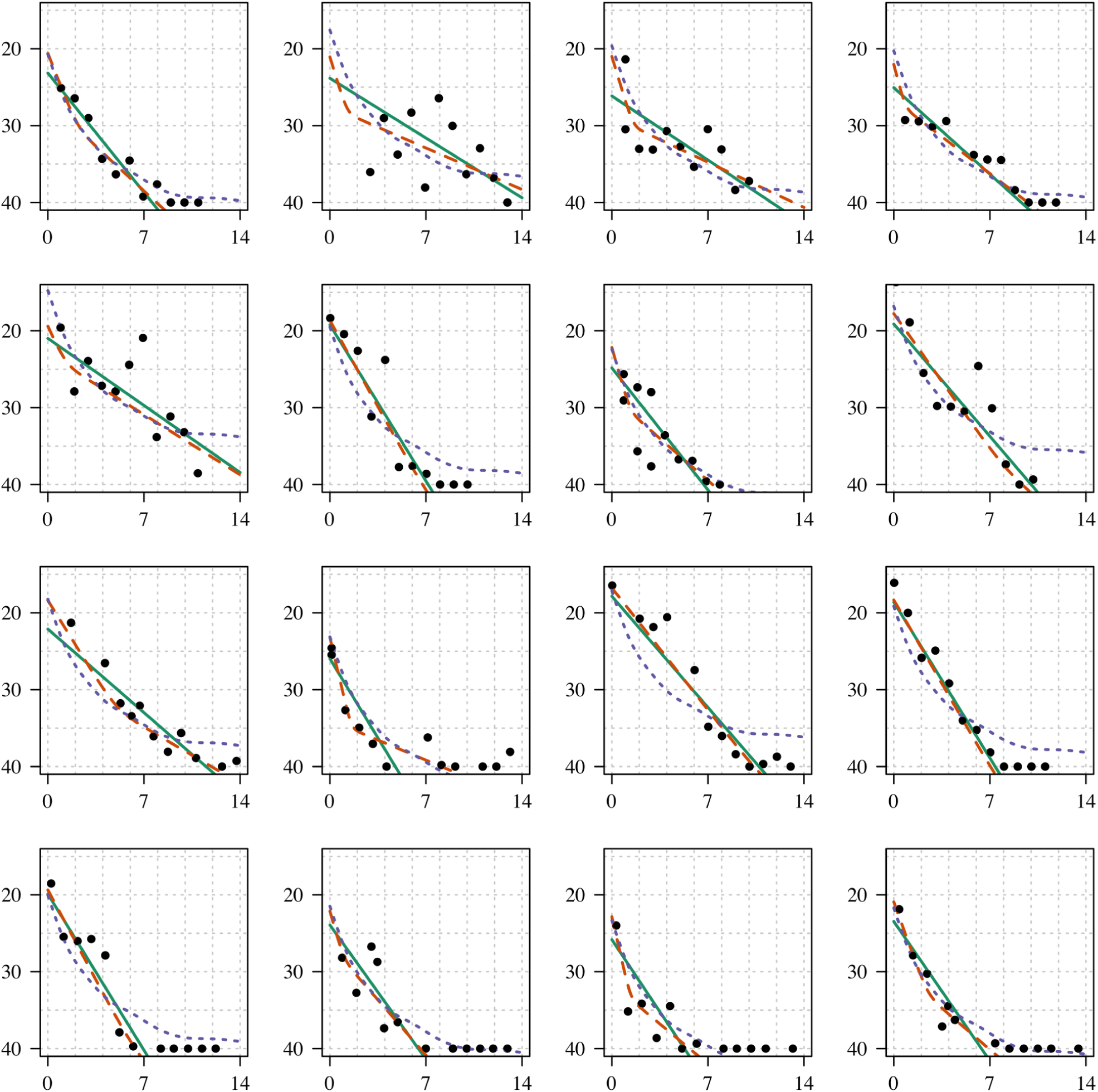
Individual model fits for infections with at least 10 CT values (panel 2). The x-axis is the time since peak viral load, the y-axis the CT value. The thick line shows the log-linear fit; the dashed line shows the bi-exponential fit.

**Figure S3:**
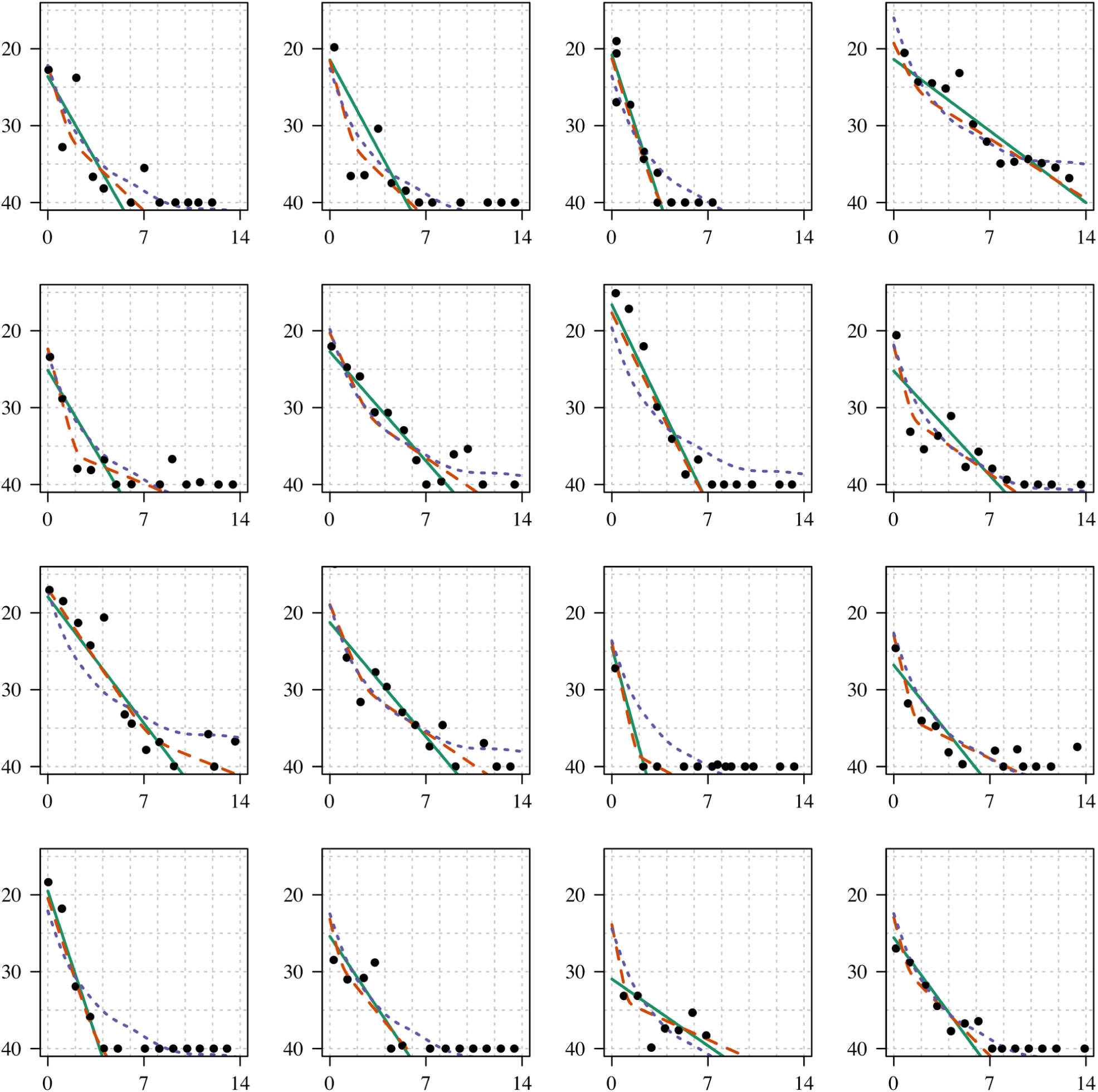
Individual model fits for infections with at least 10 CT values (panel 3). The x-axis is the time since peak viral load, the y-axis the CT value. The thick line shows the log-linear fit; the dashed line shows the bi-exponential fit.

**Figure S4:**
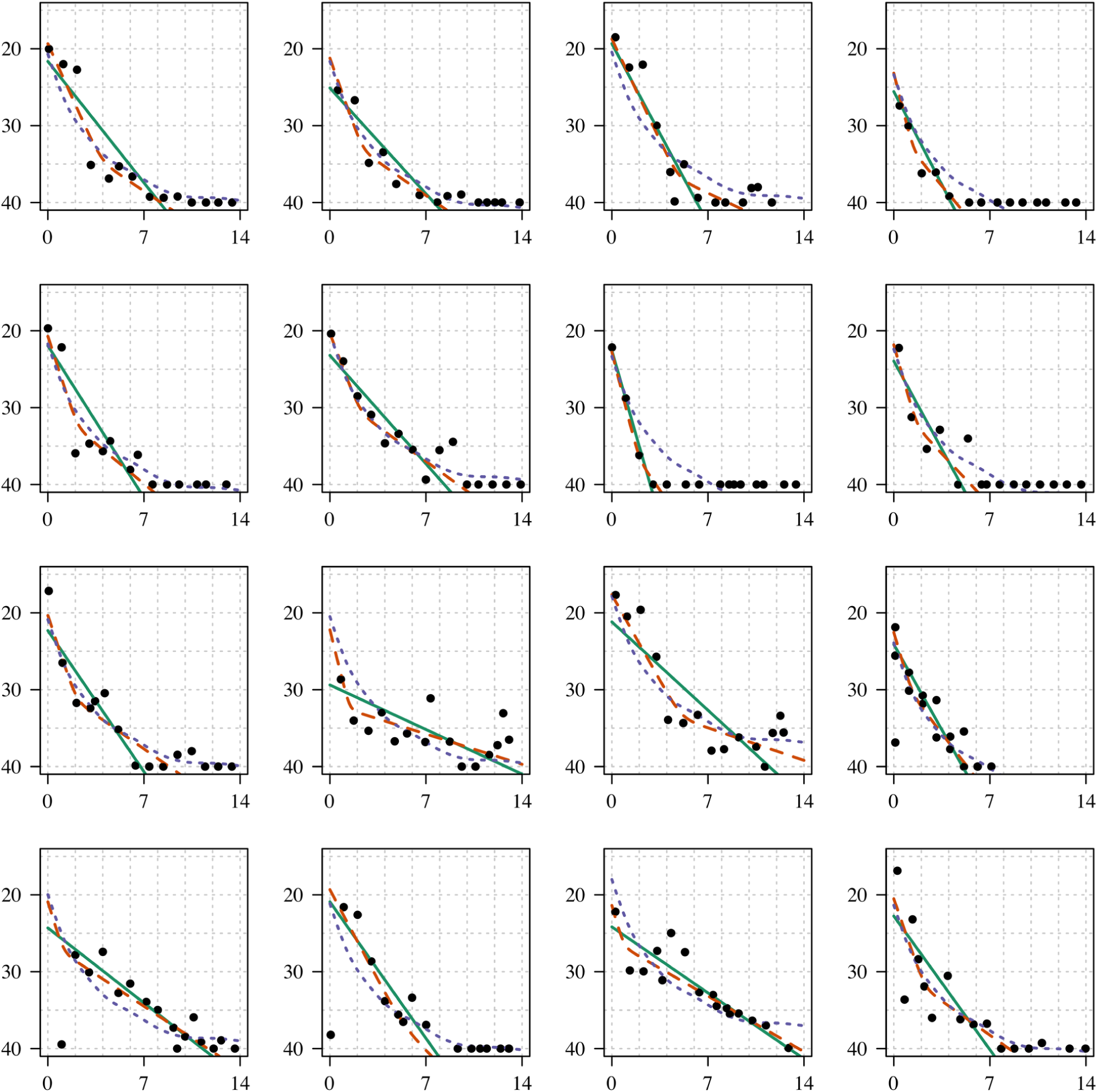
Individual model fits for infections with at least 10 CT values (panel 4). The x-axis is the time since peak viral load, the y-axis the CT value. The thick line shows the log-linear fit; the dashed line shows the bi-exponential fit.

**Figure S5:**
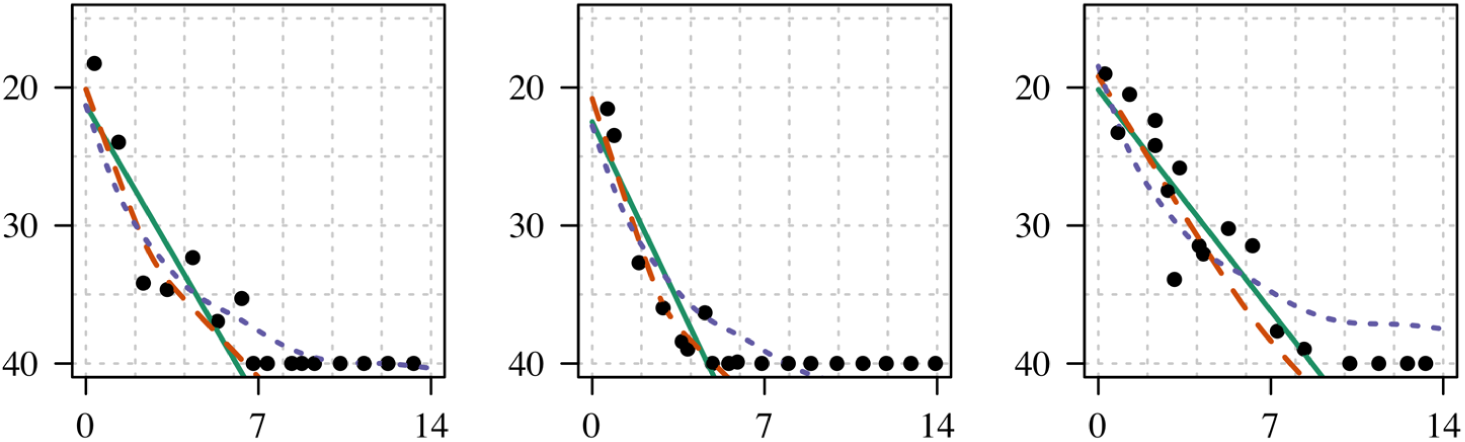
Individual model fits for infections with at least 10 CT values (panel 5). The x-axis is the time since peak viral load, the y-axis the CT value. The thick line shows the log-linear fit; the dashed line shows the bi-exponential fit.

**Figure S6:**
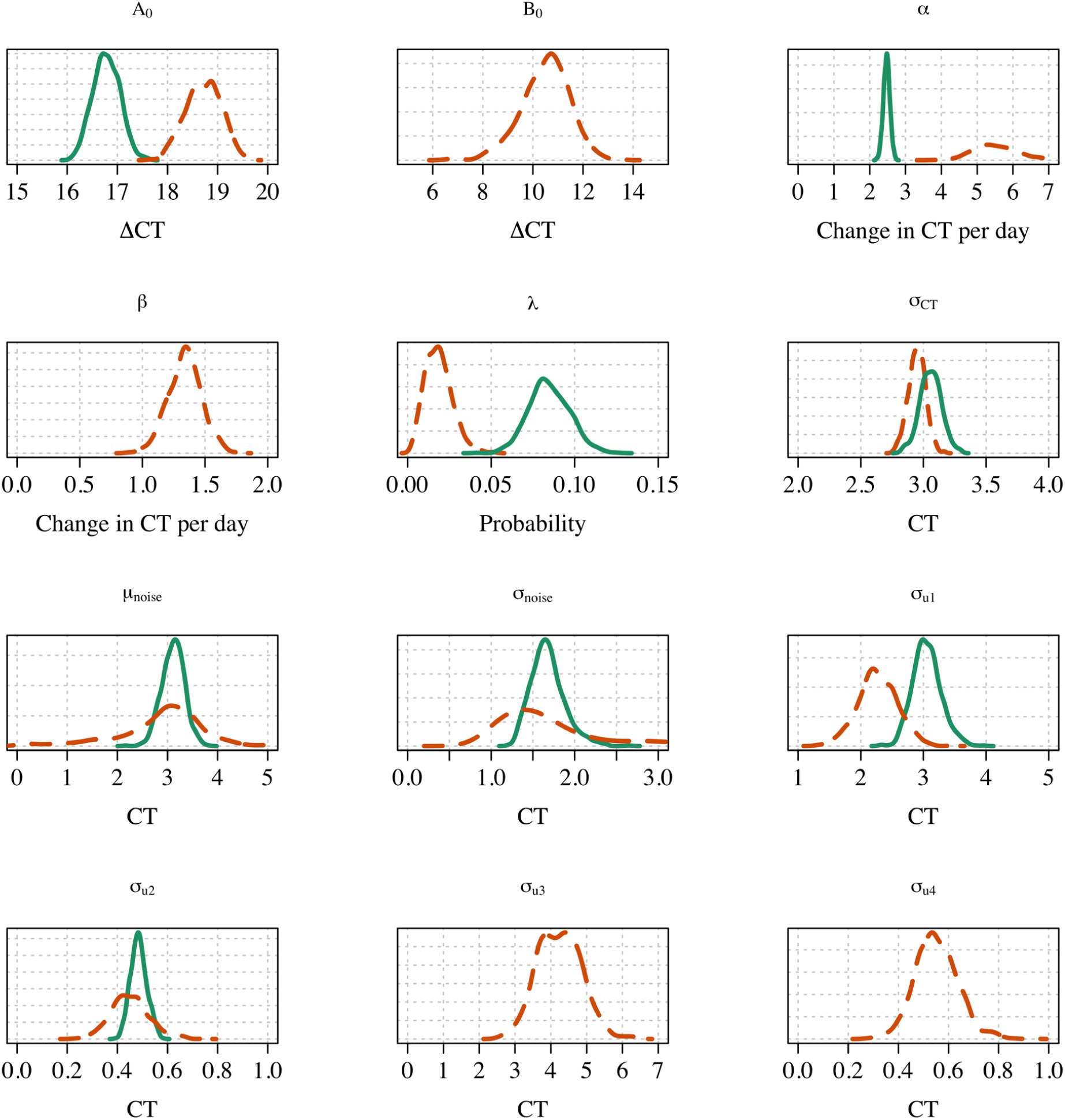
Posterior distributions for the parameters in model 1 (green thick lines) and model 2 (orange dashed lines). The parameters *σ*_*uk*_ for *k* = 1, .., 4 represent the standard deviation around the random effects *θ*_*i,k*_, respectively. *B*_0_, *β, σ*_*u*3_, *σ*_*u*4_ are parameters for model 2 only.

